# Gene expression overlap between neuropsychiatric disorders

**DOI:** 10.1101/2023.11.15.23298563

**Authors:** Alana Castro Panzenhagen, Alexsander Alves-Teixeira, Martina Schroeder Wissmann, Carolina Saibro Girardi, Lucas Santos, Alexandre Kleber Silveira, Daniel Pens Gelain, José Cláudio Fonseca Moreira

## Abstract

Common diseases result from a mix of genetic and environmental factors, often involving inflammation. Complex traits like diabetes and psychiatric disorders are polygenic, influenced by many genetic variants. The omnigenic model suggests all expressed genes can impact disease-related genes. This study examines blood transcriptomic variations in psychiatric and neurological disorders to understand mRNA expression profiles and address field discrepancies. Animal models are explored for similar gene expressions. This study extensively searched GEO DataSets and ArrayExpress databases, identifying gene expression profiles associated with neuropsychiatric disorders. From GEO, 10,359 samples were found, with 30 series (1,897 samples) in the qualitative synthesis, revealing 1,364 differentially expressed genes in Schizophrenia, 134 in Bipolar Disorder, 11 in Autism Spectrum Disorder, and 2,784 in Alzheimer’s Disorder. Comparisons with GWAS studies unveiled overlaps, with 81 genes for SCZ, two for BD, and 135 for ALZ. Notably, 441 genes were shared between ALZ and SCZ. Enrichment analyses indicated associations with signalling pathways. In animal models, 2,360 series were identified, with 175 in the qualitative synthesis, resulting in a meta-analysis focusing on ALZ with hippocampus tissue, revealing 14 consistently differentially expressed genes. Four overlapped with human data (ALOX5AP, P2RY13, RGS10, SH3GL1). These findings contribute to understanding shared and unique molecular signatures across neuropsychiatric disorders, bridging insights between human and animal models. The study efficiently identifies and tests consistent differentially expressed genes in psychiatric and neurological disorders, focusing on blood transcriptomes. Compared to transcriptome-wide or proteome-wide association studies, this approach analyses transcripts directly from individuals with disorders, offering real-world predictive capability. Shared genes between disorders suggest common molecular pathways, emphasizing the need for interdisciplinary approaches in understanding and treating psychiatric disorders. Limitations include sample characterization and the peripheral marker focus. Further investigations, including functional assays, are crucial for validation and extending these findings.

## Introduction

Many common human diseases result from a complex interplay between genetic and environmental factors, which can jointly contribute to the development of specific traits (1,2). These diseases, whether related to the central nervous system (CNS) or not, often involve inflammation, suggesting potential systemic causal or consequential effects (3–5). In the case of neurodegenerative conditions like Parkinson’s and Alzheimer’s, this association has led to the coining of the term “inflammaging” (6–8).

Conditions like diabetes, hypertension, psychiatric disorders, and even continuous traits such as body mass index (BMI) and height fall under the umbrella of complex traits. These traits are influenced by multiple genetic variants, making them polygenic and inherently intricate (1–9). This concept was partially formulated by Fisher in 1919 through his “infinitesimal model,” which posits that the contribution of each genetic variant diminishes as the number of associated genes increases, resulting in a normally distributed phenotype (10,11).

More recently, researchers have proposed that complex traits are not just polygenic but also “omnigenic.” The omnigenic model posits that all genes expressed in disease-relevant cells can affect core disease-related genes due to highly interconnected regulatory networks. This theory suggests that a substantial portion of the heritability of these traits can be attributed to genes outside of core pathways(9). While this theory is important for understanding the origins of complex traits, it must be translated into therapeutic applications when addressing common diseases. Consequently, ongoing research to identify the primary pathways responsible for susceptibility to common diseases remains vital.

In recent years, multi-omics data has been extensively generated from various human tissues and biological levels, encompassing the genome, transcriptome, proteome, and even the microbiome. These layers of data interact intricately, posing a significant challenge to contemporary bioinformatics and biostatistics research (12–14). However, the evidence derived from different studies within each omic category remains inconclusive and lacks consistency. In epidemiology, one of the most robust and commonly used statistical methods to address similar challenges is meta-analysis, which involves the comprehensive evaluation of data following a well-conducted systematic literature review to estimate overall effect sizes (15). Given that the protein networks involved in disease regulation are primarily linked to gene expression, our study focuses on the transcriptome level. Additionally, because representative tissue samples for common diseases are scarce due to high heterogeneity and complexity, the investigation of peripheral factors becomes imperative. This is particularly crucial for brain disorders, and considering potential systemic effects can provide valuable insights. Therefore, we have chosen to investigate transcriptomic variations in the most readily available peripheral tissue: blood, aiming to reconcile inconsistencies across the field.

This study aims to consolidate existing evidence on transcriptomic gene expression levels in blood samples, comparing individuals with psychiatric and neurological disorders. Through this approach, we seek to gain a comprehensive understanding of the mRNA expression profiles associated with complex traits, with the goal of resolving major discrepancies in the field. We also sought for the animal models of these disorders in order to understand if they present any similar differentially expressed genes.

## Methods and Analysis

For this study report, we have adhered to the guidelines outlined in the Preferred Reporting Items for Systematic Reviews and Meta-Analyses (PRISMA) (15).

### Eligibility Criteria

We incorporated studies that compare gene expression levels in blood samples from individuals with psychiatric and neurological disorders to those from healthy control groups. Included in our selection are experimental studies that have quantified gene expression using mRNA measurement techniques such as microarray or RNA-sequencing. We exclusively considered studies that feature a well-defined control or resilience group. Our criteria encompass randomized clinical trials (RCTs), cohort studies, and case-control studies, with the stipulation that they include a control group not subjected to specific, unique treatments. The studies should have collected blood samples (including leukocytes, lymphocytes, peripheral blood mononuclear cells (PBMCs)) from patients diagnosed with the respective disorder, as well as from healthy (or resilient) control individuals. We included all diagnostic approaches, regardless of the diagnostic manual or tool used. There were no restrictions on publication date, language, methodological quality, age, sex, or ethnicity of study participants.

We applied the following exclusion criteria: i) studies comprising only genetically related individuals (e.g., family-based studies); ii) studies lacking original data; iii) studies with cases displaying unique comorbidities; iv) studies devoid of healthy (or resilient) control groups; v) studies labeling remitters as healthy controls; vi) studies where blood samples underwent any form of ex vivo treatment before microarray analysis or RNA-sequencing; and vii) studies in which all case samples received a specific type of drug not categorized as “treatment as usual,” such as the predominant prescription drug or class of drugs.

For the animal models, the criteria were as follows:

#### Inclusion criteria

study involving rats (rat, rats, Rattus norvegicus) or mice (mouse, mice, Mus musculus); animals must serve as models (or be used as models in other studies) for the specific disorder/disease (behavioral model, knockout, lineage, etc.); presence of controls without the disorder/disease (no behavioral induction, wildtype, control lineage, etc.) or resilient animals (in this case, note in the observation!); gene expression / transcriptomic method for coding mRNA (array, microarray, RNA-Seq, RNA sequencing…) in any tissue/cell type.

#### Exclusion criteria

studies where all animal groups were treated with any drug or treatment technique (e.g., deep brain stimulation), preventing comparison between the treated and untreated model and control; studies exclusively focused on non-coding RNA, miRNA, siRNA, methylation, genome variation, genome binding…; studies conducted solely with cell cultures.

### Search and Study Identification

To identify relevant studies, we conducted an online search using two distinct electronic databases: Gene Expression Omnibus (GEO) (https://www.ncbi.nlm.nih.gov/gds) and ArrayExpress (https://www.ebi.ac.uk/arrayexpress/). We did not employ search filters.

Our search strategies encompassed subject headings for each specific disorder/disease, research involving human subjects, and blood-related research:

1. Schizophrenia: ((“schizophrenia”[mesh] OR “SCZ”) AND (“humans”[mesh] OR “Homo sapiens”[Organism]) AND (“blood”[mesh])).
2. Major depressive disorder: ((“depressive disorder”[mesh] OR “depression”[mesh] OR “depressive disorder, major”[mesh] OR “MDD” OR depress*) AND (“humans”[mesh] OR “Homo sapiens”[Organism]) AND (“blood”[mesh])).
3. ADHD: ((“attention deficit disorder with hyperactivity”[mesh] OR attention-deficit/hyperactivity disorder OR “ADHD” OR inattent* OR hyperact* OR impulsiv* OR “attention deficit”) AND (“blood”[mesh]) AND (“humans”[mesh] OR “Homo sapiens”[Organism])).
4. ALZ: ((“alzheimer disease”[mesh] OR alzheimer*) AND (“humans”[mesh] OR “Homo sapiens”[Organism]) AND (“blood”[mesh])).
5. BD: ((“bipolar disorder”[mesh] OR bipolar) AND (humans[mesh] OR “Homo sapiens”[Organism]) AND (“blood”[mesh])).
6. ASD: ((“autistic disorder”[mesh] OR “ASD” OR “autism spectrum disorder”[mesh]) AND (humans[mesh] OR “Homo sapiens”[Organism]) AND (“blood”[mesh])).

For the animal models the strategies were the following:

((“schizophrenia”[mesh] OR “SCZ”) AND (“Models, Animal”[mesh] OR “Rats”[mesh] OR “Mice”[mesh]))

((“depressive disorder”[mesh] OR “depression”[mesh] OR “depressive disorder, major”[mesh] OR “MDD” OR depress*) AND (“Models, Animal”[mesh] OR “Rats”[mesh] OR “Mice”[mesh]))

((“attention deficit disorder with hyperactivity”[mesh] OR attention-deficit/hyperactivity disorder OR “ADHD” OR inattent* OR hyperact* OR impulsiv* OR “attention deficit”) AND (“Models, Animal”[mesh] OR “Rats”[mesh] OR “Mice”[mesh]))

((“autistic disorder”[mesh] OR “ASD” OR “autism spectrum disorder”[mesh]) AND (“Models, Animal”[mesh] OR “Rats”[mesh] OR “Mice”[mesh]))

((“alzheimer disease”[mesh] OR alzheimer*) AND (“Models, Animal”[mesh] OR “Rats”[mesh] OR “Mice”[mesh]))

### Study Selection

The study selection process was conducted in two stages. Initially, we evaluated titles for inclusion, followed by a thorough review of detailed information from GEO and ArrayExpress. Both stages involved a minimum of two independent reviewers (ACP, ATT, MSW, CSG, LS, or AKT). Any disagreements were resolved through consultation with a third reviewer (ACP or JCFM).

### Data Collection

Descriptive data from each study were extracted independently by at least two reviewers (ACP, ATT, MSW, CSG, LS, or AKT), with discrepancies resolved by a third reviewer (ACP or JCFM). Data encompassed study descriptions (sample size, mean age, standard deviation, gender distribution, ethnicity, country/region of origin, data collection date, transcriptome analysis platform used, diagnostic manual/tool, and medication or psychotherapy details). Transcriptomic outcome data, including raw and/or normalized gene expression data were retrieved through the GEOquery R package [21] or direct downloads from GEO and/or ArrayExpress websites.

### Outcome Measures

Our primary outcome measure was mRNA levels, assessed through microarray or RNA-sequencing methods. These measures are typically available in full in online repositories, allowing access to raw data.

### Data Synthesis

The dataset distributions and sample quality evaluations were performed as indicated by GEO. Since not all microarray platforms had the same exact coverage, we excluded the genes that were not present in at least 85% of the studies. We imputed the remaining values with the mean from the pooled sample. All meta-analyses comprised at least 20,000 mapped genes. Gene expression data was aggregated into a single variable for each disorder using the ImaGEO shiny app, using a random effects approach and 10% missing variables allowed. Differentially expressed genes were computed collectively. We have also explored gene enrichment analyses using the FUMA tool, with at least 10 genes belonging to each set.

## Results

The search for studies in the GEO DataSets (https://www.ncbi.nlm.nih.gov/gds/) and ArrayExpress (https://www.ebi.ac.uk/arrayexpress/) databases was conducted without language or date restrictions (including studies from the inception of the database up to June 30, 2019) in accordance with the search strategies described.

In the GEO database, a total of 10,359 samples from 324 series (gene expression series - datasets, typically corresponding to a single study, except for rare duplicates) were found, of which 30 series, comprising 1,897 samples, were included in the qualitative synthesis. The inclusion process is illustrated in Figure 1. The search for studies in the ArrayExpress database did not yield any studies different from those already found through GEO.

**Figure 1.**
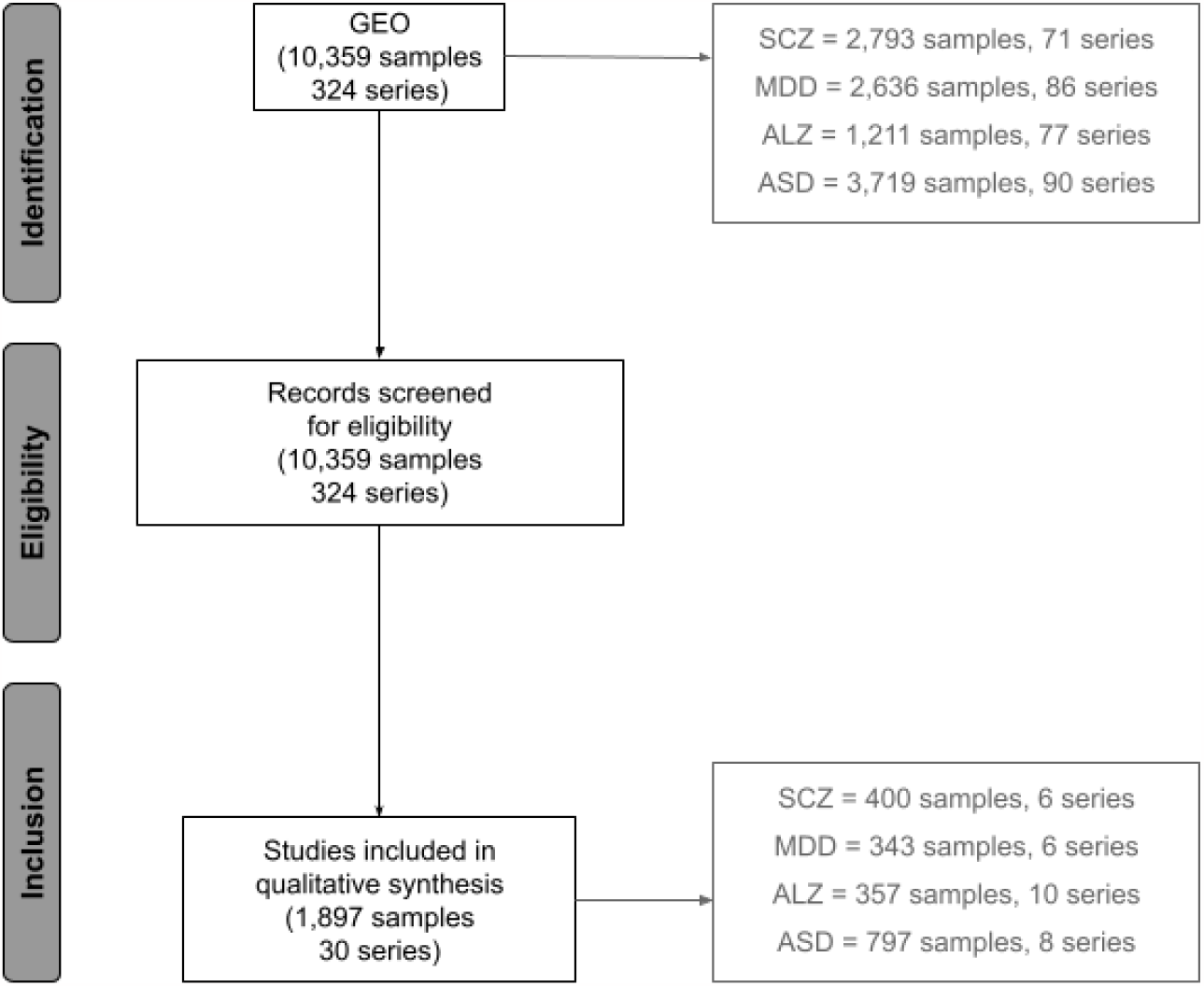
Flowchart of the study inclusion process from the GEO DataSets database for clinical studies. SCZ = Schizophrenia, MDD = Major depressive disorder, ALZ = Alzheimer’s disease, ASD = Autism spectrum disorder.

We found 1,364 consistently differentially expressed genes in Schizophrenia, 134 in Bipolar Disorder, 11 in Autism Spectrum Disorder, and 2,784 in Alzheimer’s Disorder. Figure 2A depicts the overall methodology of ImaGEO, while the first 1,000 genes associated with each disorder are represented in heatmaps (Figure 2B-E). Not all series were possible to meta-analyse mainly because of different built for microarrays, making it difficult to join the data.

**Figure 2.**
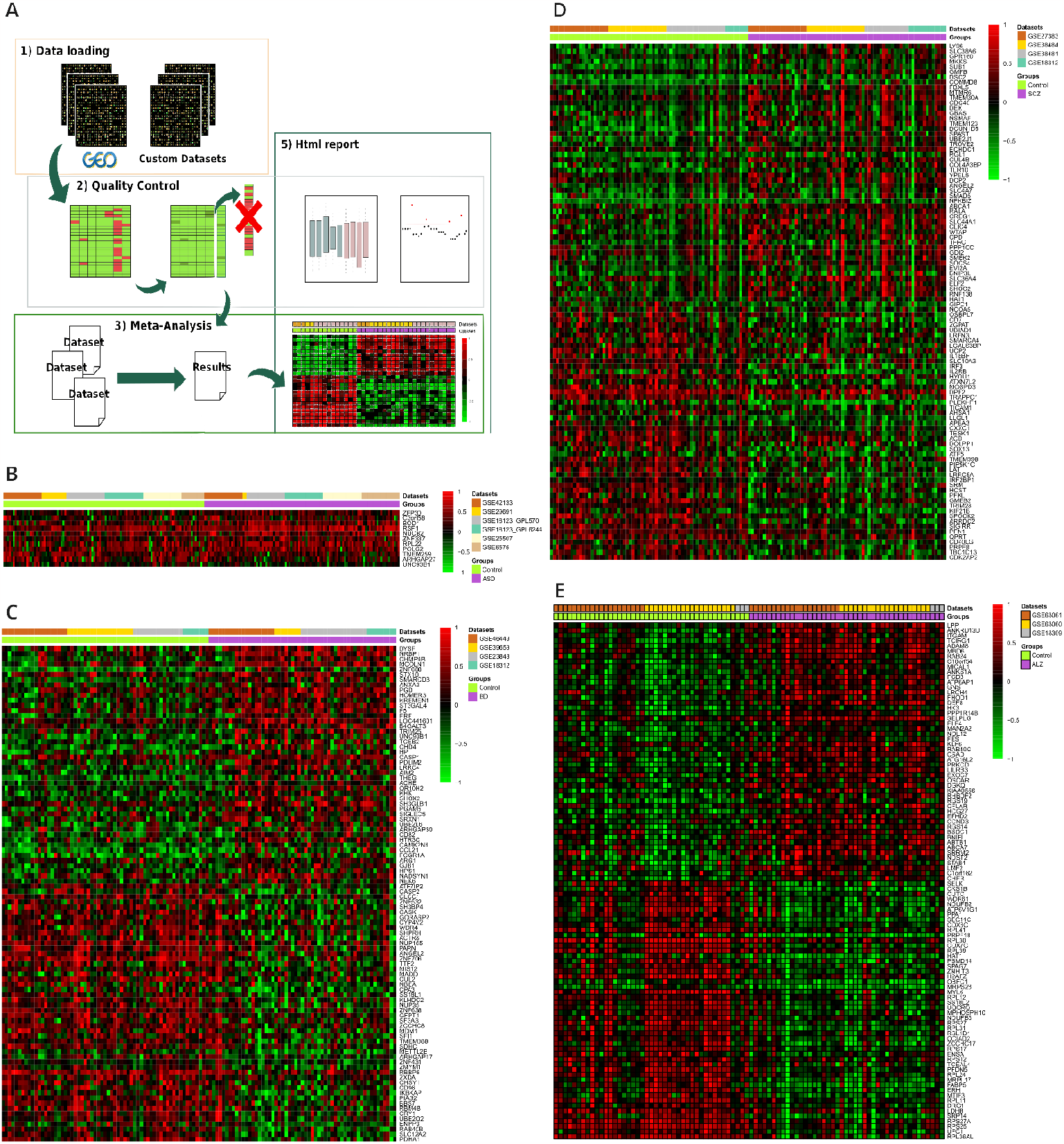
Gene expression meta-analysis results for the first 1000 genes. (A) The overall methodology of ImaGEO shinny app. Gene expression heatmaps representing the associations found in Autism Spectrum Disorder (B), Bipolar Disorder (C), Schizophrenia (D), and Alzheimer’s disease (E).

When comparing the differentially expressed genes found here with those significantly associated with outcomes in GWAS studies (data from the GWAS Catalog - https://www.ebi.ac.uk/gwas/), we found 81 overlapping for SCZ, two for BD, and 135 for ALZ (Supplementary file 1).

We also found that the differentially expressed genes overlap to some extend in between disorders (Figure 3A), with the biggest overlap being between ALZ and SCZ (441 genes). We have investigated these genes further through enrichment analyses. SCZ genes seem to be associated with signalling pathways, proteolysis, endocytosis, and cell cycle (Figure 3B). ALZ genes also seem to be associated with signalling pathways and the cell cycle, but mainly with the ribosome, oxidative phosporilation, Huntington’s disease, and cancer (Figure 3C). When we look at the intersection between ALZ and SCZ genes we see they are mainly associated with Toll-like receptor signalling, proteolysis, endocytosis and cancer pathways (Figure 3D).

**Figure 3.**
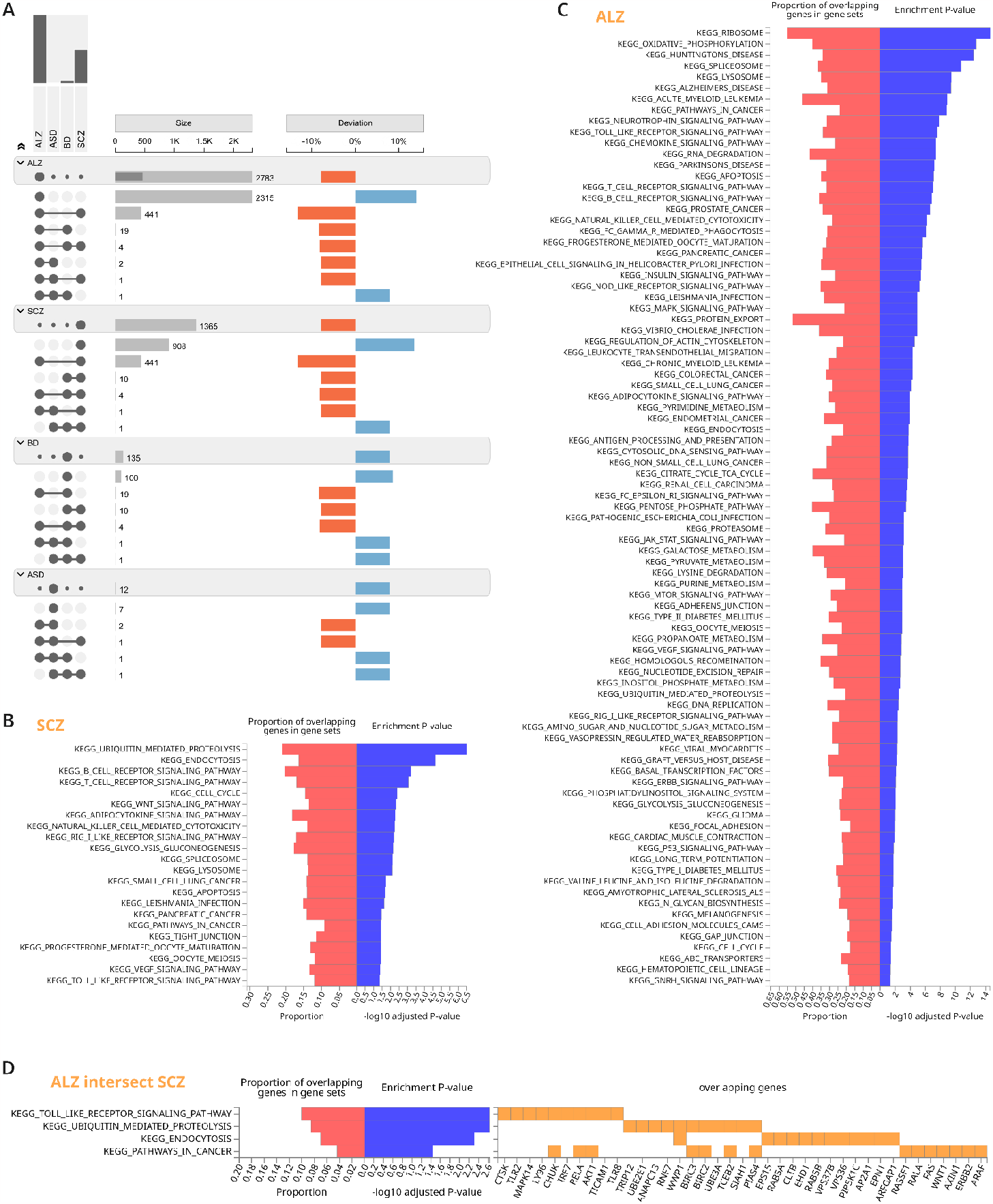
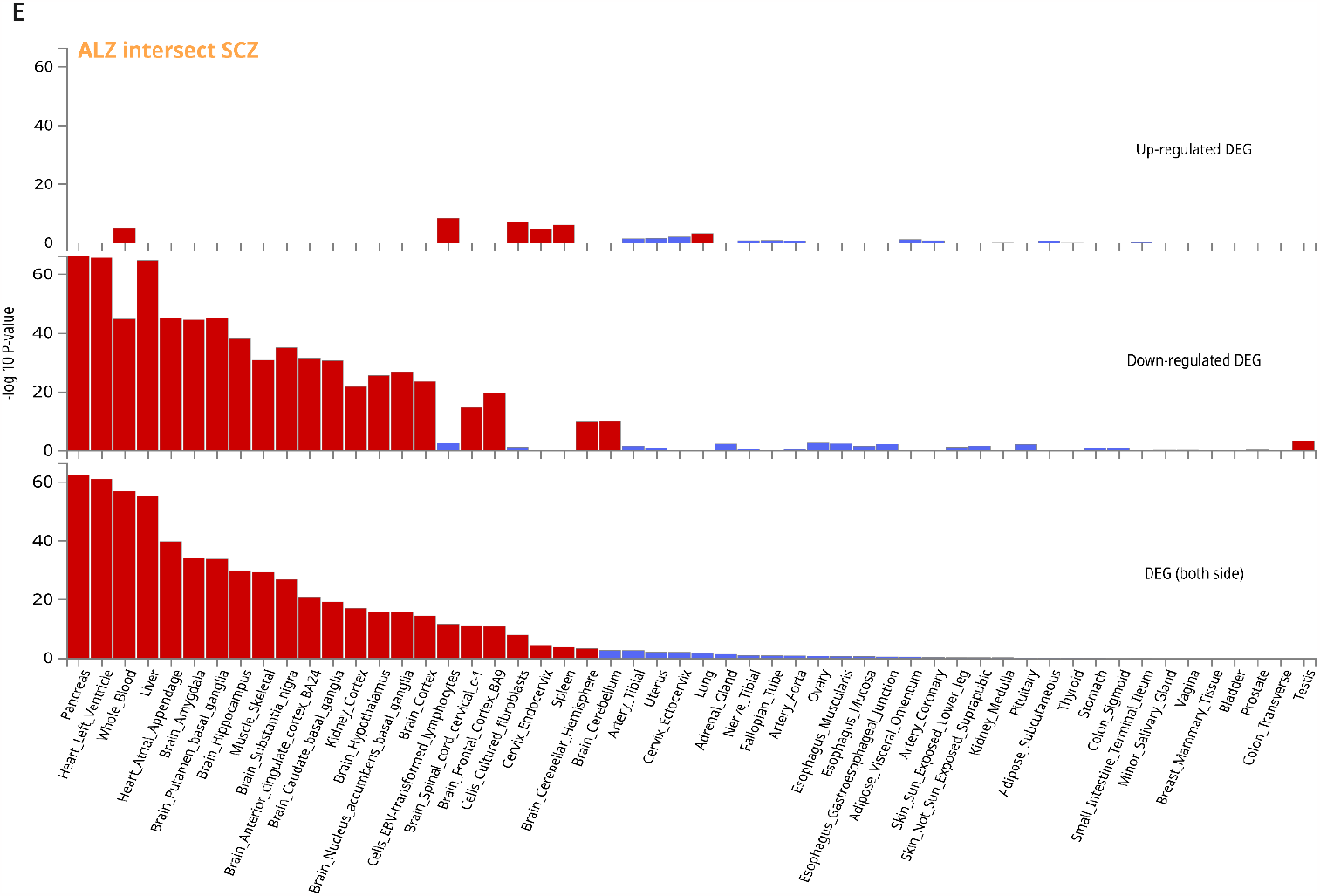
Enrichment analysis of Schizophrenia and Alzheimer’s associated genes. All enrichment analyses are corrected by FDR and only genets with more than 10 genes were considered. A) The overlap between all disorders tested in an UpSet style plot. B) Enrichment analysis of Schizophrenia-associated genes in the Kyoto Encyclopaedia of Genes and Genomes (KEGG). C) Enrichment analysis of Alzheimer’s-associated genes in KEGG. D) Enrichment analysis of genes overlapping between Alzheimer’s and Schizophrenia. E) Tissue-specific expression of genes in the overlap between Alzheimer’s and Schizophrenia; red bars represent significant associations < 0.05 after FDR correction. ALZ = Alzheimer’s disease, ASD = Autism Spectrum Disorder, BD = Bipolar Disorder, SCZ = Schizophrenia.

Regarding the animal models, we identified 2,360 series and included 175 in our qualitative synthesis, comprising 2,040 samples (Figure 4). However, we were only able to conduct a meta-analysis with the ALZ group with hippocampus tissue due to a lack of data or comparable tissues. We found 14 genes consistently differentially expressed (Figure 5). Four of those were also found in our meta-analysis with human data (ALOX5AP, P2RY13, RGS10, SH3GL1).

**Figure 4.**
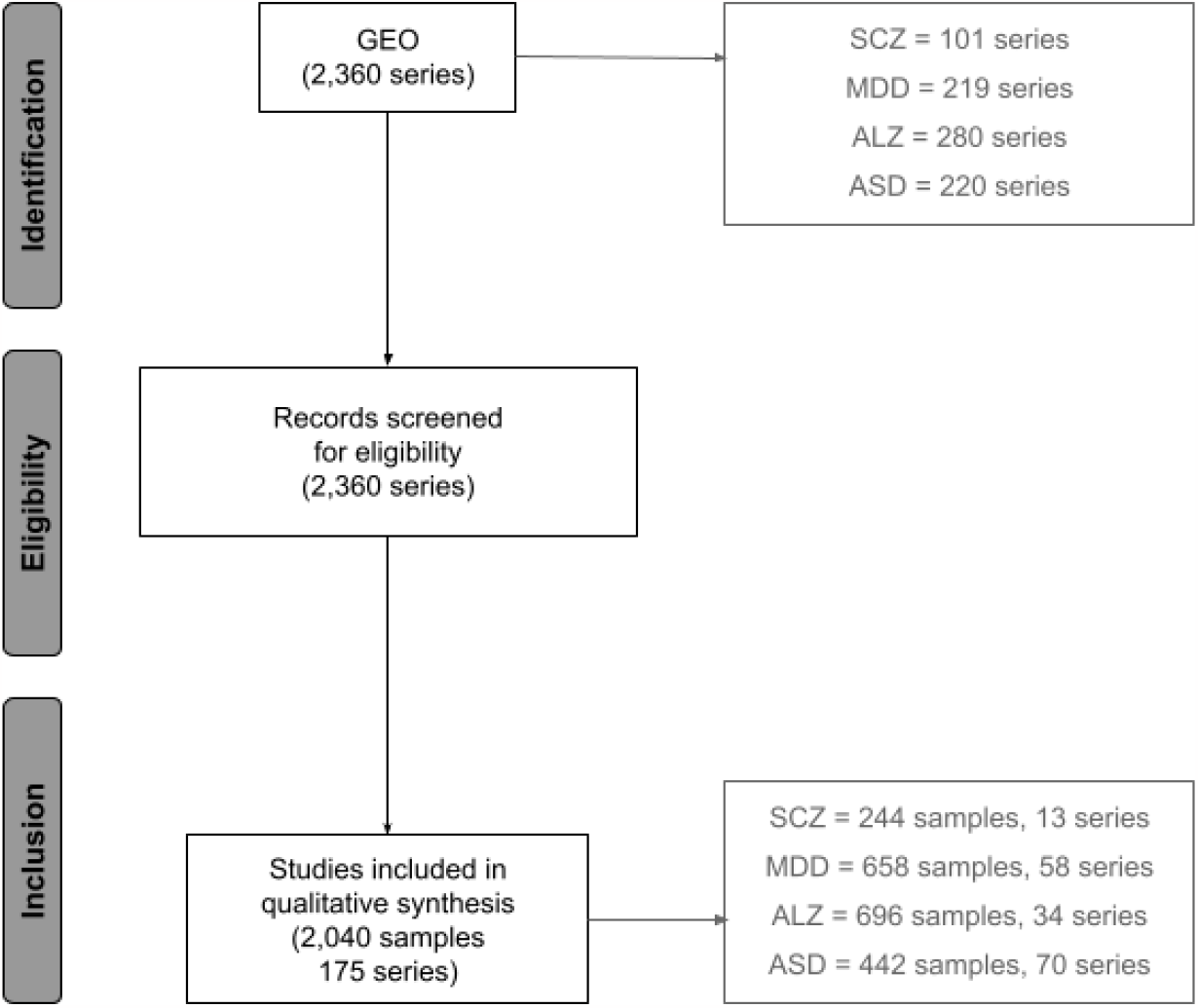
Flowchart of the study inclusion process from the GEO DataSets database for animal model studies. SCZ = Schizophrenia, MDD = Major depressive disorder, ALZ = Alzheimer’s disease, ASD = Autism spectrum disorder.

**Figure 5.**
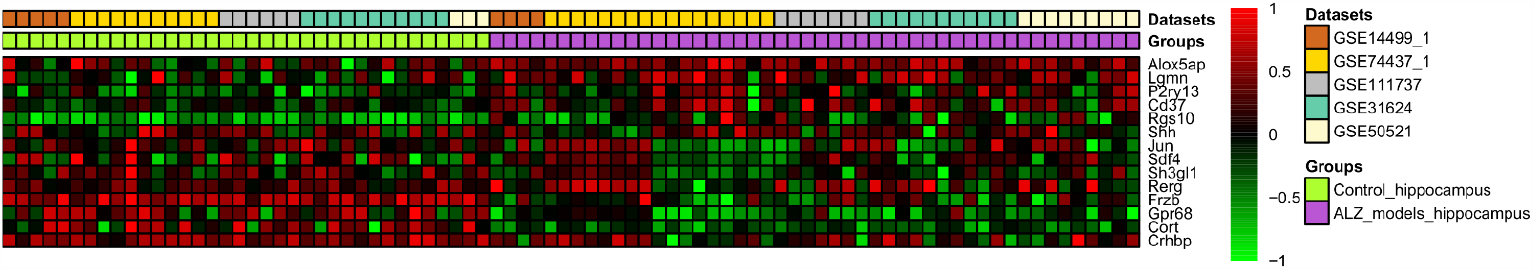
Gene expression meta-analysis results in heatmaps representing the associations found in Alzheimer’s disease animal models hippocampal tissue.

All genes described here and the qualitative synthesis of studies can be found in Supplementary file 1.

## Discussion

The methodology described demonstrates an efficient technique for identifying and testing the consistency of differentially expressed genes across multiple studies with a similar design. The differentially expressed genes that were found still need to be individually further investigated for us to better interpret these results. However, the focus of the present study is on the overlap of markers between disorders and the similarities found in relation to GWAS results.

One of the main differences from our study to the ones that perform transcriptome-wide association (TWA) or proteome-wide association (PWA) is that we are meta-analysing the transcripts found directly in individuals with the disorders. The disadvantage of this approach is that it is highly improbable that we would be able to identify the whole set of causal genes or variants, since it is a cross-sectional freeze of the current characteristics, after years with the disorder. On the other hand, the advantage is that it carries more predictive capability in real-world samples, as we can identify the consequences of the disorder on the transcriptome. Future studies should focus on analyses that are carried out at different stages of disorder development, for example near the onset, or after different treatments. This could potentially lead us to better biomarkers that could be used in the clinic. Another advantage of this approach is that we can now compare what is found at the individual level with the predicted changes in TWAS and PWAS studies to better dissect what has been causal or consequential to the condition.

In a bid to contextualize our results, we compared the differentially expressed genes with those significantly associated with outcomes in GWAS studies, drawing data from the GWAS Catalog (16). Notably, 81 genes were overlapped in SCZ, two in BD, and 135 in ALZ. It shows that some of the genes associated in the transcriptome were also associated in GWA studies, and we would argue, most likely have to do with the development of the disorder.

A recent study by Wingo and colleagues, from 2022 compared the shared mechanisms across major psychiatric and neurodegenerative diseases by mapping TWAS and PWAS predictions from summary statistics of GWAS (17). They tried to infer the most likely causal genes for the overlap between disorders. They have found 13 different genes that were in the intersection between psychiatric and neurodegenerative diseases, from which we see two were also in our SCZ-ALZ overlap (HSDL1 and STXBP3). We could interpret as if those genes are part of a shared molecular pathophysiology that should be taken into account in early identification and treatment.

A particularly intriguing aspect of our investigation lies in the exploration of shared genes among the different psychiatric disorders. Subsequent enrichment analyses shed light on the functional implications of these shared genes. SCZ-associated genes were found to be linked with signaling pathways, proteolysis, endocytosis, and the cell cycle. On the other hand, ALZ-associated genes exhibited associations with signalling pathways, the cell cycle, the ribosome, oxidative phosphorylation, Huntington’s disease, and cancer. The intersection of ALZ and SCZ genes was particularly noteworthy, with associations identified in Toll-like receptor signalling, proteolysis, endocytosis, and cancer pathways. The overlap between SCZ and ALZ has not been often noted in GWA studies (18,19) and it seems to be more prevalent when we investigate consequences and markers of the disorders in contrast to its genetic causes. In neuroimaging, for example, that is also true, in which several structural and functional changes are similar in between SCZ and ALZ (20–22). The environmental interactions might be at play here, in which similar environmental factors or endophenotypes, such as cognitive decline, depressive behaviour, and others could be responsible by the shared biology that we encounter in these neuropsychiatric disorders. It is also possible that this relationship between SCZ and ALZ may still become evident in GWAS as larger samples are collected. Nevertheless, another possibility is that GWAS studies may not be sufficient to investigate the heritability of these disorders, emphasizing the importance of interdisciplinary approaches, as presented here. When we look at the overlap between ALZ and SCZ in terms of where these genes are expressed, we found that, apart from some constitutive expressions in the pancreas, heart, and liver, they were also found highly expressed in brain amygdala, putamen, hippocampus, the substantia nigra, and the caudate; suggesting key changes in CNS functions.

Some limitations should be taken into account when interpreting our results, especially: i) that a meta-analysis is just as good as its original studies, and we were limited as to characterization of samples and sample sizes; ii) that using peripheral markers, although non-invasive and good for predictions, hinders our ability to derive interpretations about the CNS and what are the changes that occur in the brain.

Our findings not only deepen our understanding of the intricate molecular landscape of psychiatric disorders but also hint at potential shared pathways and mechanisms across seemingly distinct conditions. Further investigations, including functional assays and validation studies, will be crucial to validate and extend these initial observations. The integration of diverse datasets and advanced analytical approaches continues to unveil the complex nature of psychiatric disorders, offering new avenues for targeted therapeutic interventions and precision medicine strategies.

## Data Availability

All data produced in the present study are available upon reasonable request to the authors

## References

1. Quintana-Murci L. Understanding rare and common diseases in the context of human evolution. Genome Biology 2016 17:1 [Internet]. 2016 Nov 7 [cited 2023 Nov 15];17(1):1–14. Available from: https://genomebiology.biomedcentral.com/articles/10.1186/s13059-016-1093-y

2. Furlong LI. Human diseases through the lens of network biology. Trends Genet [Internet]. 2013 Mar [cited 2023 Nov 15];29(3):150–9. Available from: https://pubmed.ncbi.nlm.nih.gov/23219555/

3. Rocha EM, De Miranda B, Sanders LH. Alpha-synuclein: Pathology, mitochondrial dysfunction and neuroinflammation in Parkinson’s disease. Neurobiol Dis. 2018 Jan 1;109:249–57.

4. Sampson TR, Debelius JW, Thron T, Janssen S, Shastri GG, Ilhan ZE, et al. Gut Microbiota Regulate Motor Deficits and Neuroinflammation in a Model of Parkinson’s Disease. Cell [Internet]. 2016 Dec 1 [cited 2023 Nov 15];167(6):1469–1480.e12. Available from: https://pubmed.ncbi.nlm.nih.gov/27912057/

5. Michopoulos V, Powers A, Gillespie CF, Ressler KJ, Jovanovic T. Inflammation in Fear- and Anxiety-Based Disorders: PTSD, GAD, and Beyond. Neuropsychopharmacology [Internet]. 2017 Jan 1 [cited 2023 Nov 15];42(1):254–70. Available from: https://pubmed.ncbi.nlm.nih.gov/27510423/

6. McGeer PL, Rogers J, McGeer EG. Inflammation, Antiinflammatory Agents, and Alzheimer’s Disease: The Last 22 Years. Journal of Alzheimer’s Disease. 2016;54(3):853–7.

7. Franceschi C, Garagnani P, Vitale G, Capri M, Salvioli S. Inflammaging and ‘Garb-aging.’ Trends in Endocrinology & Metabolism. 2017 Mar 1;28(3):199–212.

8. Xia S, Zhang X, Zheng S, Khanabdali R, Kalionis B, Wu J, et al. An Update on Inflamm-Aging: Mechanisms, Prevention, and Treatment. J Immunol Res [Internet]. 2016 [cited 2023 Nov 15];2016. Available from: https://pubmed.ncbi.nlm.nih.gov/27493973/

9. Boyle EA, Li YI, Pritchard JK. An expanded view of complex traits: from polygenic to omnigenic. Cell [Internet]. 2017 Jun 6 [cited 2023 Nov 15];169(7):1177. Available from: /pmc/articles/PMC5536862/

10. Barton NH, Etheridge AM, Véber A. The infinitesimal model: Definition, derivation, and implications. Theor Popul Biol [Internet]. 2017 Dec 1 [cited 2023 Nov 15];118:50–73. Available from: https://pubmed.ncbi.nlm.nih.gov/28709925/

11. Fisher RA. XV.—The Correlation between Relatives on the Supposition of Mendelian Inheritance. Earth Environ Sci Trans R Soc Edinb [Internet]. 1919 [cited 2023 Nov 15];52(2):399–433. Available from: https://www.cambridge.org/core/journals/earth-and-environmental-science-transactions-of-royal-society-of-edinburgh/article/abs/xvthe-correlation-between-relatives-on-the-supposition-of-mendelian-inheritance/A60675052E0FB78C561F66C670BC75DE

12. Huang S, Chaudhary K, Garmire LX. More is better: Recent progress in multi-omics data integration methods. Front Genet. 2017 Jun 16;8(JUN):268903.

13. Vasaikar S V., Straub P, Wang J, Zhang B. LinkedOmics: analyzing multi-omics data within and across 32 cancer types. Nucleic Acids Res [Internet]. 2018 Jan 1 [cited 2023 Nov 15];46(D1):D956–63. Available from: https://pubmed.ncbi.nlm.nih.gov/29136207/

14. Hawe JS, Theis FJ, Heinig M. Inferring interaction networks from multi-omics data. Front Genet. 2019 Jun 12;10(JUN):440512.

15. Page MJ, McKenzie JE, Bossuyt PM, Boutron I, Hoffmann TC, Mulrow CD, et al. The PRISMA 2020 statement: an updated guideline for reporting systematic reviews. Syst Rev [Internet]. 2021 Dec 1 [cited 2023 Nov 15];10(1):1–11. Available from: https://systematicreviewsjournal.biomedcentral.com/articles/10.1186/s13643-021-01626-4

16. Buniello A, Macarthur JAL, Cerezo M, Harris LW, Hayhurst J, Malangone C, et al. The NHGRIEBI GWAS Catalog of published genome-wide association studies, targeted arrays and summary statistics 2019. Nucleic Acids Res [Internet]. 2019 Jan 8 [cited 2023 Nov 15];47(D1):1005–12. Available from: 10.1093/nar/gky1120

17. Wingo TS, Liu Y, Gerasimov ES, Vattathil SM, Wynne ME, Liu J, et al. Shared mechanisms across the major psychiatric and neurodegenerative diseases. Nature Communications 2022 13:1 [Internet]. 2022 Jul 26 [cited 2023 Nov 15];13(1):1–19. Available from: https://www.nature.com/articles/s41467-022-31873-5

18. Smajlagic D, Zayats T, Bekkehus M, Hellard S Le. Genome-wide association studies in psychiatry: Current perspectives. European Psychiatry [Internet]. 2021 Apr [cited 2023 Nov 15];64(S1):70–1. Available from: 10.1192/j.eurpsy.2021.219

19. Mallard TT, Grotzinger AD, Smoller JW. Examining the shared etiology of psychopathology with genome-wide association studies. Physiol Rev [Internet]. 2023 Apr 1 [cited 2023 Nov 15];103(2):1645–65. Available from: 10.1152/physrev.00016.2022

20. Demirhan A. The effect of feature selection on multivariate pattern analysis of structural brain MR images. Physica medica : PM : an international journal devoted to the applications of physics to medicine and biology : official journal of the Italian Association of Biomedical Physics [Internet]. 2018 Mar 1 [cited 2023 Nov 15];47:103–11. Available from: 10.1016/j.ejmp.2018.03.002

21. Noor MBT, Zenia NZ, Kaiser MS, Mamun S Al, Mahmud M. Application of deep learning in detecting neurological disorders from magnetic resonance images: a survey on the detection of Alzheimer’s disease, Parkinson’s disease and schizophrenia. Brain Inform [Internet]. 2020 Dec 1 [cited 2023 Nov 15];7(1). Available from: 10.1186/s40708-020-00112-2

22. Hassanzadeh R, Abrol A, Calhoun V. Classification of Schizophrenia and Alzheimer’s Disease using Resting-State Functional Network Connectivity. 2022 IEEE-EMBS International Conference on Biomedical and Health Informatics (BHI) [Internet]. 2022 [cited 2023 Nov 15];01–4. Available from: 10.1109/BHI56158.2022.9926797

